# Assessing the efficacy of behaviourally informed invitation messaging in increasing attendance at the NHS Targeted Lung Health Check: A randomised experimental study

**DOI:** 10.64898/2026.04.12.26350693

**Authors:** Xiaozhou Tan, Martin Danka, Szymon Urbanski, Pam Kitsawat, Terence J. McElvaney, Shyma Jundi, Lucy Porter, Chiara Gericke

## Abstract

**Background:** Lung cancer screening can reduce lung cancer mortality through early detection, but uptake of the NHS Targeted Lung Health Check (TLHC) programme remains low. Behaviourally informed invitation messages have been proposed as a low-cost approach to increase attendance, but evidence of their effectiveness in lung cancer screening is mixed. Few intervention studies used evidence-based behaviour change frameworks, and rarely tailored invitation strategies to empirically identified barriers and enablers.

**Methods:** In an online experiment, 3,274 adults aged 55-74 years and with a history of smoking were randomised to see one of four behaviourally informed invitation messages or a control message. Participants then rated their intention to attend a TLHC appointment, and selected barriers and enablers to attending from a pre-defined list, which were classified according to the Theoretical Domains Framework. Invitation messages were mapped to Behaviour Change Techniques using the Theory and Techniques Tool. Message conditions were compared on intention to attend TLHC using bootstrapped ANOVA followed by pairwise comparisons. Exploratory counterfactual mediation analyses examined the role of fear in intention to attend.

**Results:** Behaviourally informed invitation messages did not meaningfully increase intention to attend TLHC compared with the control message. While a GP-endorsed message showed a small potential benefit relative to the other conditions, this finding was not robust after adjustment for multiple comparisons. Participants most frequently reported barriers related to *Emotion* (particularly fear), *Social Influence*, and *Knowledge*, while *Beliefs about Consequences* emerged as the primary enabler of attendance. Only around half of reported barriers and enablers were addressed by the invitation messages. Exploratory analyses found that fear was associated with lower intention to attend a TLHC appointment, yet none of the behaviourally informed messages appeared to reduce fear compared to the control message.

**Conclusions:** Improving lung cancer screening uptake will likely require invitation messages that directly address emotional concerns, particularly fear, alongside credible recommendations. These findings highlight the importance of systematically aligning invitation message content with empirically identified behavioural influences when designing scalable interventions to improve lung cancer screening uptake.

**Contributions to the literature:**

- Attendance at the NHS Targeted Lung Health Check remains low despite being a UK policy priority. This study provides evidence to inform invitation strategies for ongoing and future screening programmes.
- Behavioural economics frameworks are widely used in health messaging but lack systematic methods for linking behavioural influences with intervention content and evaluation.
- This study demonstrates how Behaviour Change frameworks can diagnose why behaviourally informed messages may fail, by assessing alignment between intervention content and population-specific barriers and enablers.
- Findings suggest that messaging interventions targeting lung cancer screening attendance may have limited impact when they do not address emotional and social influences.

## 1. Introduction

Smoking is a leading cause of lung cancer mortality, accounting for approximately 18.4% of all cancer deaths worldwide (Bray et al., 2018). Evidence from large-scale trials demonstrates that low-dose computed tomography (LDCT) screening can reduce lung cancer mortality by around 20% through earlier detection (Field et al., 2021). Survival outcomes for lung cancer vary by stage at diagnosis, with five-year survival rates of approximately 65% for Stage 1 disease, compared to 40%, 15%, and 5% for Stages 2, 3, and 4 respectively (Cancer Research UK, 2022). Accordingly, the National Health Service (NHS) launched the Targeted Lung Health Check (TLHC) programme in 2019 to facilitate earlier diagnosis among individuals at higher risk of lung cancer. It was rebranded to the NHS Lung Cancer Screening (LCS) programme (UK National Screening Committee, 2025) in January 2025, but the term TLHC is retained throughout for consistency with the study period and existing literature. The programme is offered to people aged 55 and over who are current or former smokers that reside in the United Kingdom (National Health Service, 2022). Individuals eligible for the TLHC programme are invited to attend an initial lung health assessment, which may be completed in-person, telephone, or online. Those assessed as eligible are subsequently scheduled for an LDCT scan at their nearest NHS hospital.

The effectiveness of LDCT screening depends on uptake, yet participation remains low (on average, 35%) and is socially patterned, with lower response and attendance rates observed among those at highest risk of lung cancer, including current smokers and individuals from lower socio-economic backgrounds (Bartlett et al., 2020; Crosbie et al., 2019; Dickson et al., 2023). Improving attendance is a stated policy priority in the UK, with a recent House of Commons briefing identifying the TLHC as a key mechanism for earlier detection in cancers with poorer survival outcomes (House of Commons Library, 2026). Because attendance is initiated through an invitation, improving the effectiveness of invitation communications offers a promising route for increasing screening uptake and reducing health inequalities.

Behaviour change frameworks provide structured approaches for understanding influences of health-related behaviours such as screening attendance, and for designing interventions that target these influences. The Theoretical Domains Framework (TDF) synthesises constructs from 33 behavioural theories into 14 domains that can be used to classify barriers and enablers based on their underlying mechanism of action (Cane et al., 2012; Atkins et al., 2017). The Behaviour Change Wheel (BCW) builds on this by systematically linking identified influences on appropriate intervention functions and Behaviour Change Techniques (BCTs) (Michie et al., 2011, 2013). This approach can be used both in the development of novel behaviour change interventions, and in the evaluation of existing interventions (e.g., by assessing the theoretical fit between identified influences on a behaviour and the intervention content that has been delivered to change that behaviour). Evidence indicates that theory-based interventions are more effective than those lacking an explicit theoretical foundation (Michie & Prestwich, 2010). The Theory and Techniques Tool (TaTT) supports this approach by providing a matrix linking BCTs to the TDF domains (alongside additional mechanisms of action) based on a triangulation of prior evidence and expert consensus (Michie et al., 2021; Carey et al., 2019). This process ensures intervention content targets specific influences based on established evidence and provides a basis for evaluating both impact and implementation processes.

These frameworks have been applied to identify barriers and enablers to cancer screening uptake across multiple contexts, including lung cancer (Salman et al., 2025), cervical cancer (Stuart & D’Lima, 2022), and breast cancer (Huf et al., 2025). Lung cancer screening adherence is influenced by interacting psychological, social, and contextual factors; using the TDF to synthesise influences from 73 studies, Salman and colleagues (2025) identified ‘Emotion’ as the most frequently reported barrier, particularly fear of a cancer diagnosis, often linked to fatalistic beliefs, and fear of the screening procedure. Barriers were also evident within ‘Environmental Context and Resources’, including appointment inconvenience and perceived financial or logistical costs, and within ‘Knowledge’, where limited understanding of screening purpose, benefits, eligibility, and the need for screening in the absence of symptoms reduced perceived relevance and uptake. Pahwa et al. (2025) further highlighted stigma as an additional emotional barrier, with (ex-)smokers reporting the fear of stigma of their smoking history.

In contrast, ‘Emotion’ may also operate as an enabler. Olson et al. (2022) found that fear and worry about cancer outcomes motivated screening participation, prompting participants to seek reassurance through a negative result or, where a positive diagnosis felt inevitable, to confirm it early enough for treatment or psychological preparation. Related to this, ‘Beliefs about Consequences’ was identified as a prominent enabler (Salman et al., 2025), with beliefs in the benefits of early detection, including improved treatment prospects and extended survival, strengthening motivation to attend (Schiffelbein et al., 2020). In addition, ‘Social Influences’ were frequently reported as enablers, whereby clinician recommendation and encouragement from trusted others increased screening attendance (Salman et al., 2025).

Based on the BCW intervention development framework, interventions to improve lung cancer screening attendance should target TDF domains as identified above. Existing interventions to increase lung cancer screening uptake vary in design and theoretical grounding. A systematic review of 13 randomised trials found that few interventions explicitly used validated evidence-based behaviour change frameworks, and rarely tailored invitation strategies to empirically identified barriers and enablers (Yang et al., 2025). The most consistent positive effects were observed in higher-intensity interventions involving direct interaction with healthcare professionals, typically combining education about screening benefits with practical barrier resolution (Yang et al., 2025). In contrast, low-cost messaging interventions (e.g., invitation letters) showed modest and inconsistent effects. Moreover, some studies lacked clear theoretical grounding altogether (e.g., Adler et al., 2024), and even where behavioural theory was referenced, it was rarely preceded by systematic assessment of population-specific enablers and barriers prior to message design (Yang et al., 2025). Consequently, how theoretically informed interventions influence attendance remains unclear.

A broader body of research from other cancer screening programmes and the NHS Health Check indicates that behaviourally informed messaging can improve attendance (Uy et al., 2017; Sallis et al., 2019). A review of behavioural science-based interventions across cancer screening programmes reported improvements in uptake ranging from 4% to 63%, although effect sizes varied widely by context (Uy et al., 2017). These interventions commonly utilised frameworks such as MINDSPACE (Dolan et al., 2010) and concepts such as “nudge” (Thaler and Sunstein, 2008). Techniques associated with improved uptake include personalisation, prompting individuals to form explicit intentions or plans to attend, emphasising the potential consequences of non-attendance, highlighting social norms, and providing clear scheduling information such as deadlines (Hirst et al., 2016; Hagoel et al., 2016).

Nevertheless, the effectiveness of these techniques is inconsistent across settings (Uy et al., 2017), and their added benefit over standard invitations is not always observed. In summary, while barriers and enablers to lung cancer screening attendance have been well characterised (Salman et al., 2025), and messaging interventions have shown some promise in other cancer screening contexts (Uy et al., 2017; Hirst et al., 2016; Hagoel et al., 2016), evidence from lung cancer screening interventions remains mixed. Many of these interventions draw on behavioural science frameworks (e.g., MINDSPACE), but they do not provide methods for linking identified barriers and enablers to intervention techniques. Evaluations also rarely assess whether intervention techniques align with the influences most relevant to the target population (Yang et al., 2025). Consequently, whether existing invitation messages correctly address the key influences on lung cancer screening attendance remains unclear.

## The present study

To inform efforts to improve TLHC uptake, the NHS conducted a randomised experiment in October 2023 comparing four behaviourally informed invitation messages with a control message based on current invitation letter wording. The intervention messages were informed by behavioural economics frameworks (Hallsworth et al., 2015), prior NHS Health Check studies (Sallis et al., 2016; 2019), and expert consensus, but were not developed using the Behaviour Change Wheel approach.

This study presents the first published analysis of the invitation-message data from this trial, evaluating the intervention messages in terms of both their impact on intentions to attend and their alignment with participant-reported barriers and enablers. Specifically, it examines the extent to which behaviour change techniques (BCTs) within the messages address the Theoretical Domains Framework (TDF) domains characterising participants’ reported influences on attendance. In doing so, the study provides a systematic, theory-informed evaluation of invitation messages and addresses a key policy priority for improving lung cancer screening uptake.

Accordingly, the study addresses the following research questions:

1. Are behaviourally informed invitation messages more effective than a non-behaviourally informed message in increasing participants’ intention to attend the TLHC?
2. What barriers and enablers do participants report in relation to attending lung cancer screening (using the TDF)?
3. To what extent did the intervention messages address the key barriers and enablers reported by participants?
4. Exploratory: What role does emotion, specifically fear, play in the relationship between invitation messages and participants’ intention to attend the TLHC?

## 2. Methods

### 2.1 Design

This research presents the first published analysis of the invitation-message data from a between-subjects randomised experimental study conducted by the NHS England Behavioural Science Unit in collaboration with the NHS Cancer Programme Team. The broader study aimed to identify ways to increase participation in the TLHC programme among adults aged 55–74 years with a history of smoking. See Appendix A for details of the full study.

### 2.2 Ethical Approval

All participants provided informed consent prior to participation in the study. Ethical approval was obtained through NHS England’s Behavioural Science Unit internal governance procedures. Regarding this secondary data analysis, the project was registered with UCL Data Protection (reference number “Z6364106/2024/07/89 health research”) in line with UCL’s Data Protection Policy. The data is publicly available on Open Science Framework, via https://osf.io/qvjwy/) and no further ethical approval was required for this analysis.

### 2.3 Recruitment

Participants were recruited via a market research agency (Bilendi), the NHS App, and NHS Vaccines volunteer research panels. Eligibility criteria included being aged 55 to 74 years, residing in England, and having a history of smoking (current or former smokers). Data collection took place between 15 September 2023 and 2 October 2023.

### 2.4 Original Study Procedure

Participants completed the study online via Qualtrics. Respondents were first screened for eligibility. Individuals who did not meet the eligibility criteria were excluded and thanked.

Participants were assigned a unique anonymous identifier generated by Qualtrics and allocated to one of five invitation-message conditions. Randomisation was implemented after eligibility screening, entirely within the Qualtrics survey platform, using the built-in “Randomizer” function with Evenly Present Elements enabled. This algorithm combines a simple random allocation with balancing: the platform tracks per-arm counts and nudges assignment towards under-represented arms to maintain an approximately 1:1:1:1:1 ratio. No blocking or stratification was used. Allocation concealment was maintained because the randomisation algorithm was embedded in the survey flow and not visible to participants or researchers.

After allocation to a condition, participants viewed their assigned invitation message, then completed the primary outcome measure (self-reported likelihood of attending the TLHC), followed by ratings of emotional responses, reasons for attending and not attending the TLHC, and demographic information. The full survey took approximately 13 minutes to complete.

### 2.5 Measures and Materials

#### 2.5.1 Invitation messages

All invitation messages shared a common introductory opening. They differed in that each additionally included one of four types of behavioural messaging (see Table 1 for the intervention message content):

1. **Gain-frame message:** Emphasised the positive outcomes of early detection and the success of treatment when screening is attended.
2. **Messenger message:** Used endorsement from a credible health professional (the participant’s GP) to enhance motivation and trust.
3. **Status quo message:** Framed attendance as timely and personally relevant, used prompts and cues to reduce hesitation and procrastination.
4. **Challenging assumptions message:** Sought to address misconceptions that attending screening burdens the NHS, reframing non-attendance as the costlier choice.

**Table 1.**
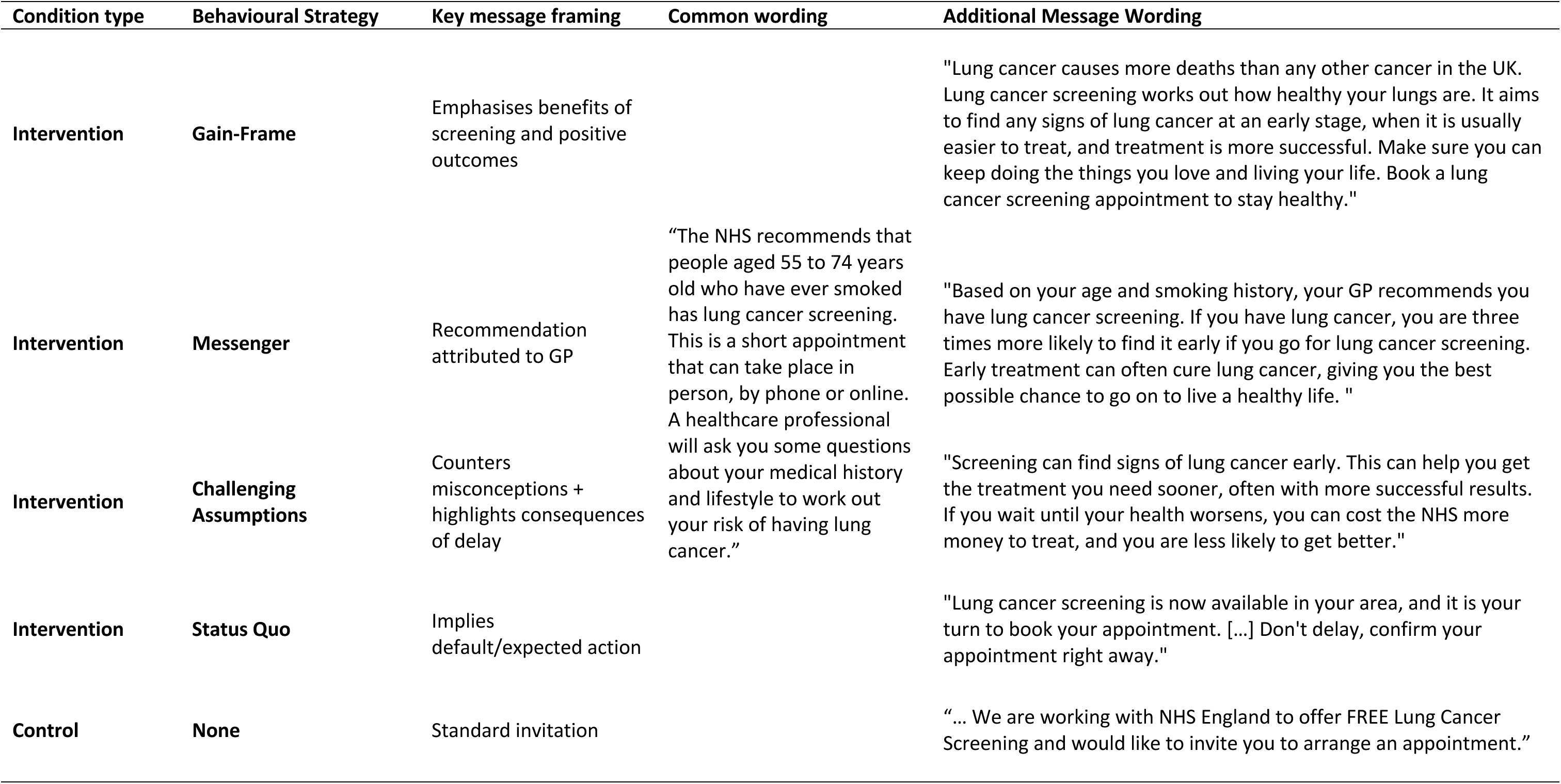
Message contents.

The Control condition also used the common opening, followed by wording drawn from current TLHC invitation letters. It provided factual information and instructions on booking and attending an appointment but did not include additional behavioural components. Appendix B contains the full details of each message.

#### 2.5.2 Outcomes

After seeing one of the five messages, participants were asked “Based on what you have read, on a scale of 1 (not likely at all) to 7 (very likely), how likely would you be to take part?” Participants then selected up to three reasons why they *would* attend the TLHC from a list of eight options, and up to three reasons they would not attend the TLHC from a list of eleven options. The full list of reasons is provided in Appendix C.

#### 2.5.3 Mediators

Participants were asked “How did this message make you feel? You can choose up to three emotions” from a predefined list of ten, presented in a randomised order. The list of emotions contained four positive (encouraged, interested, confident, grateful), two neutral (uninterested, nothing), and four negative (annoyed, confused, scared, nervous). The development of this list was guided by established classifications of positive and negative discrete emotions in social psychology, particularly Fredrickson’s (2013) work on positive emotions and Izard’s (1997) Differential Emotions Theory.

#### 2.5.4 Demographic and health-related questions

A series of demographic and health-related questions were asked, namely: age, gender, ethnicity, education level, employment status, accommodation status, smoking status, self-rated general health, health literacy, and whether they had a long-term lung condition.

### 2.6 Data analysis

We used both qualitative and quantitative methods to address the research questions. All quantitative analyses were conducted in R v4.4.3, and qualitative data analysis was conducted in Microsoft Excel.

#### 2.6.1 Effect of message condition on intention (RQ1)

The effect of message condition on intentions to attend a TLHC appointment was analysed using one-way ANOVA. Because the outcome variable was measured on a Likert scale, the p-value was obtained using studentised bootstrap performed on the Welch-type F statistic with 5,000 resamples, which is robust to departures from normality and homoskedasticity. Where significant effects were observed, pairwise comparisons were conducted using Welch’s t-tests with Bonferroni correction and bootstrapped confidence intervals and p-values. Bootstrapped ANOVAs were implemented using the WRS2 R package v1.1-6 (Mair & Wilcox, 2020) and bootstrapping for the t-tests was done using the MKinfer package v1.3 (Kohl, 2025).

#### 2.6.2 Barriers and enablers to attendance (RQ2)

To explore participants’ reported enablers and barriers to attending the TLHC, multiple choice responses describing reasons for attending or not attending were deductively coded to the TDF (Cane et al., 2012). Coding was conducted by XT and reviewed by LP and CG, with discrepancies resolved through discussion until consensus was reached. Responses were then summarised using descriptive statistics to identify the most commonly reported barriers and enablers to attending in terms of the TDF.

#### 2.6.3 Alignment of messages with barriers and enablers (RQ3)

To examine whether the invitation messages addressed the barriers and enablers identified in RQ2, each invitation message was coded for BCTs using the BCTTv1 (Michie et al., 2013; see Appendix D for the full list of BCTs identified in each message). The TaTT (Michie et al., 2021) was then used to assess the links between these BCTs and the relevant TDF domains identified in the analysis for RQ2.

A “link” was recorded where the TaTT indicated a confirmed or inconclusive link between the BCT and the relevant TDF domain. A “non-link” was recorded where the TaTT indicated a confirmed non-link or no evidence between the BCT and the relevant TDF domain. Links and non-links were visualised through a framework mapping diagram to illustrate which barriers and enablers were addressed by the behavioural content of the messages.

#### 2.6.4 Mediation by feeling scared or nervous (RQ4)

A counterfactual-based mediation analysis examined whether feeling scared or nervous transmitted the effect of messaging on participants’ intention to attend the TLHC. These emotions were selected because prior lung cancer screening research consistently identifies emotional concerns, including fear of diagnosis and anxiety about screening procedures, as key barriers to screening attendance (Salman et al., 2025). Other negative emotions (e.g., feeling confused or annoyed) more likely reflect informational- or acceptability-related issues, which were not frequently reported in the literature.

Because both the mediator (emotions) and the outcome (intention to attend) were measured simultaneously, temporal ordering could not be guaranteed. We considered it more plausible that the reported emotional response would precede the intention to attend but given this and other strong assumptions the mediation analyses should be regarded as exploratory.

Mediation analyses estimate direct and indirect effects: the indirect effect represents the effect of messaging on TLHC that operates through emotions, whereas the direct effect operates through other pathways. Several definitions of direct and indirect effects exist; we used the interventional approach (Didelez et al., 2006; VanderWeele et al., 2014). An overview of this approach and assumptions is provided in Nguyen et al. (2021).

Prior to mediation analyses, we estimated the exposure-mediator and mediator-outcome associations, using logistic regression models for the messaging-emotion relationships and linear regression for the emotion-intention relationships. Interventional direct and indirect effects (IDE, IIE) were estimated using a Monte Carlo simulation approach, implemented in the mediation R package v4.5.1 (Imai et al. 2010), with confidence intervals obtained using 2000 bootstrap samples. The estimated IDE and IIE contrasts compared each experimental condition against control as the reference, with all participants contributing to each contrast. Although treatment was randomised, this does not remove potential confounding of the mediator-outcome relationship. We therefore adjusted for sociodemographic (age, gender, ethnicity, socioeconomic status) and health-related (smoking status, health literacy, long-term lung condition) influences in all models.

## 3. Results

### 3.1 Participants

The study included 3,274 participants (50.5% male, 49.5% female) with a mean age of 65.1 years (SD = 5.5). Most participants did not report a long-term lung condition (79.1%) and identified as ex-smokers (71.0%). Table 2 presents the full demographic distribution across the message conditions. The randomisation process ensured that participants were evenly distributed across these conditions.

**Table 2.** Distribution of participants across the invitation message conditions by demographics factors.

| Characteristic | Control<br>N = 717<sup>1</sup> | Gain-frame<br>N = 637<sup>1</sup> | Messenger<br>N = 642<sup>1</sup> | Status quo<br>N = 630<sup>1</sup> | Challenging assumptions<br>N = 648<sup>1</sup> | Overall<br>N = 3,274<sup>1</sup> | Missing, n (%) |
| --- | --- | --- | --- | --- | --- | --- | --- |
| **Likelihood of attending** | 6.04 (1.59) | 6.04 (1.47) | 6.22 (1.28) | 6.04 (1.51) | 6.15 (1.38) | 6.09 (1.45) | 0 |
| **Feeling scared** | 34 (4.7%) | 40 (6.3%) | 50 (7.8%) | 39 (6.2%) | 42 (6.5%) | 205 (6.3%) | 0 |
| **Feeling nervous** | 118 (16.5%) | 84 (13.2%) | 116 (18.1%) | 115 (18.3%) | 116 (17.9%) | 549 (16.8%) | 0 |
| **Gender** |  |  |  |  |  |  | 10 (0.3%) |
| Male | 355 (49.6%) | 320 (50.4%) | 338 (52.8%) | 297 (47.4%) | 338 (52.2%) | 1,648 (50.5%) |  |
| Female | 361 (50.4%) | 315 (49.6%) | 302 (47.2%) | 329 (52.6%) | 309 (47.8%) | 1,616 (49.5%) |  |
| **Age (years)** | 65.1 (5.5) | 64.9 (5.4) | 65.3 (5.4) | 65.3 (5.4) | 65.3 (5.5) | 65.1 (5.5) | 0 |
| **Ethnicity** |  |  |  |  |  |  | 18 (0.5%) |
| Other | 22 (3.1%) | 19 (3.0%) | 25 (3.9%) | 25 (4.0%) | 14 (2.2%) | 105 (3.2%) |  |
| White | 692 (96.9%) | 614 (97.0%) | 613 (96.1%) | 601 (96.0%) | 631 (97.8%) | 3,151 (96.8%) |  |
| **Smoking status** |  |  |  |  |  |  | 0 |
| Ex-Smoker | 515 (71.8%) | 449 (70.5%) | 460 (71.7%) | 456 (72.4%) | 444 (68.5%) | 2,324 (71.0%) |  |
| Smoker | 202 (28.2%) | 188 (29.5%) | 182 (28.3%) | 174 (27.6%) | 204 (31.5%) | 950 (29.0%) |  |
| **Education** |  |  |  |  |  |  | 37 (1.1%) |
| University Degree | 219 (30.8%) | 190 (30.2%) | 184 (29.1%) | 202 (32.2%) | 194 (30.4%) | 989 (30.6%) |  |
| Training | 251 (35.4%) | 212 (33.7%) | 226 (35.8%) | 194 (30.9%) | 239 (37.5%) | 1,122 (34.7%) |  |
| No Degree | 240 (33.8%) | 228 (36.2%) | 222 (35.1%) | 231 (36.8%) | 205 (32.1%) | 1,126 (34.8%) |  |
| **Accommodation** |  |  |  |  |  |  | 35 (1.1%) |
| Not own | 199 (27.9%) | 177 (28.1%) | 185 (29.0%) | 167 (26.9%) | 171 (26.7%) | 899 (27.8%) |  |
| Own accom | 513 (72.1%) | 453 (71.9%) | 452 (71.0%) | 453 (73.1%) | 469 (73.3%) | 2,340 (72.2%) |  |
| **Employment status** |  |  |  |  |  |  | 10 (0.3%) |
| Unemployed | 432 (60.6%) | 375 (59.1%) | 413 (64.5%) | 402 (63.9%) | 427 (65.9%) | 2,049 (62.8%) |  |
| Employed | 258 (36.2%) | 234 (36.9%) | 207 (32.3%) | 213 (33.9%) | 194 (29.9%) | 1,106 (33.9%) |  |
| Other | 23 (3.2%) | 25 (3.9%) | 20 (3.1%) | 14 (2.2%) | 27 (4.2%) | 109 (3.3%) |  |
| **Long-term lung condition** | 153 (21.5%) | 110 (17.5%) | 142 (22.4%) | 120 (19.3%) | 119 (18.6%) | 644 (19.9%) | 40 (1.2%) |
| **Health literacy score** | 3.61 (0.66) | 3.63 (0.66) | 3.63 (0.66) | 3.64 (0.64) | 3.63 (0.65) | 3.63 (0.65) | 0 |
| **General health** |  |  |  |  |  |  | <10 (<0.3%) |
| Excellent/very good | 143 (20.0%) | 151 (23.7%) | 126 (19.6%) | 136 (21.7%) | 133 (20.6%) | 689 (21.1%) |  |
| Good | 265 (37.1%) | 215 (33.8%) | 220 (34.3%) | 218 (34.7%) | 235 (36.4%) | 1,153 (35.3%) |  |
| Fair/poor | 307 (42.9%) | 271 (42.5%) | 296 (46.1%) | 274 (43.6%) | 277 (42.9%) | 1,425 (43.6%) |  |

*Note. <sup>1</sup>n (%); Mean (SD). Percentages are computed for each variable from the total of participants with observed data for that variable.*

The sample was predominantly White (96.8%), reflecting the older age range of participants (55-74 years), in which ethnic diversity is lower than in the general population. Consequently, the number of participants from minority ethnic groups was small, limiting the ability to examine ethnic differences in responses or outcomes in this analysis. While most analyses included the entire sample, 134 (4.1%) participants were excluded from the exploratory mediation analyses due to missing data on at least one covariate, and fewer than ten participants who identified as non-binary were excluded because the small cell size precluded stable model estimation.

### 3.2 Effect of message condition on intention (RQ1)

The bootstrapped ANOVA suggested that the average self-reported likelihood of attending the TLHC differed between the message conditions (*F* = 2.53, *p* = .039). The results of the bootstrapped pairwise comparisons are presented in Table 3. On average, participants assigned to the Messenger condition reported a 0.19-point higher likelihood of attending the TLHC compared to those in the Control (95% CI [0.03, 0.34], p = .015), Gain-frame (95% CI [0.04, 0.34], p = .014), and the Status Quo conditions (95% CI [0.04, 0.34], p = .017). However, these differences did not pass the adjusted significance threshold following the Bonferroni correction (α _Bonferroni_ = 0.005). Other estimated effects were close to null, with confidence intervals crossing zero.

**Table 3.** Pairwise comparisons using bootstrapped Welch’s t-test.

| Comparisons | Mean Difference [95% CI] | p |
| --- | --- | --- |
| Messenger - Control | 0.19 [0.03, 0.34] | .015* |
| Messenger - Gain-frame | 0.19 [0.04, 0.34] | .014* |
| Messenger - Status Quo | 0.19 [0.04, 0.34] | .017* |
| Messenger - Challenge | 0.08 [-0.07, 0.23] | .274 |
| Gain-frame - Control | 0.00 [-0.17, 0.16] | .996 |
| Status Quo - Control | 0.00 [-0.16, 0.17] | .990 |
| Challenge - Control | 0.11 [-0.05, 0.27] | .170 |
| Gain-frame - Status Quo | 0.00 [-0.17, 0.16] | .956 |
| Gain-frame - Challenge | -0.11 [-0.26, 0.05] | .182 |
| Status Quo - Challenge | -0.11 [-0.26, 0.05] | .182 |

*Note. Analyses were based on a sample of 3274 participants. CI = confidence interval. \* $p < .05$*

### 3.3 Barriers and enablers to attendance

Participants responded to a multiple-choice question including 19 predefined reasons for attending or not attending a TLHC, comprising eight enablers and eleven barriers. Of these, 17 items were mapped to six of the 14 domains of the TDF. Two items could not be categorised within any TDF domain without making assumptions about their underlying mechanism of action:

“I always attend health screening when invited”

“I would not want to know if I have lung cancer”

The full list of reasons and their coding is provided in Appendix C.

The frequency of reported barriers and enablers, organised by TDF domain, is summarised in Table 4. The three most commonly reported enablers for participants to attend the TLHC were Beliefs about Consequences (e.g., “If cancer is diagnosed earlier, it is more treatable”) (N = 2657, 81.2%), Emotion (e.g., “It would be reassuring if I was told I didn’t have cancer”) (N = 1510, 46.1%), and Optimism (e.g., “It is something that I can do to take control of my health”) (N = 923, 28.2%). The three most frequently reported barriers for participants to attend the TLHC were Social Influence (e.g., “My GP has not recommended it to me”) (N = 1229, 37.5%), Emotion (e.g., “I would be scared to find out if I have cancer”) (N = 1040, 31.8%), and Knowledge (e.g., “I don’t have any symptoms”) (N = 844, 25.8%). See Appendix C for the full table including rationale for TDF coding decisions.

**Table 4.** Participants’ reported enablers and barriers coded with TDF.

| TDF Domains | n | % |
| --- | --- | --- |
| Enablers |  |  |
| Beliefs about consequences | 2657 | 81.2 |
| Social influence | 657 | 20.1 |
| Optimism | 923 | 28.2 |
| Emotion | 1510 | 46.1 |
| Environmental context and resources | 458 | 14.0 |
| N/A | 1154 | 35.2 |
| Barriers |  |  |
| Beliefs about consequences | 286 | 8.7 |
| Social influence | 1229 | 37.5 |
| Emotion | 1040 | 31.8 |
| Knowledge | 844 | 25.8 |
| Environmental context and resources | 388 | 11.9 |
| N/A | 168 | 5.1 |

*Note: N/A due to no sufficient evidence to group as any TDF domain without making assumptions.*

### 3.4 Alignment of messages with barriers and enablers (RQ3)

Overall, 57.0% of the reported enablers and barriers were addressed by the messages, covering four out of the six coded TDF domains. The unaddressed domains were Emotion and Optimism.

Figure 1 illustrates the links between TDF domains associated with participants’ reported barriers and enablers and the BCTs used in the messages.

**Figure 1.**
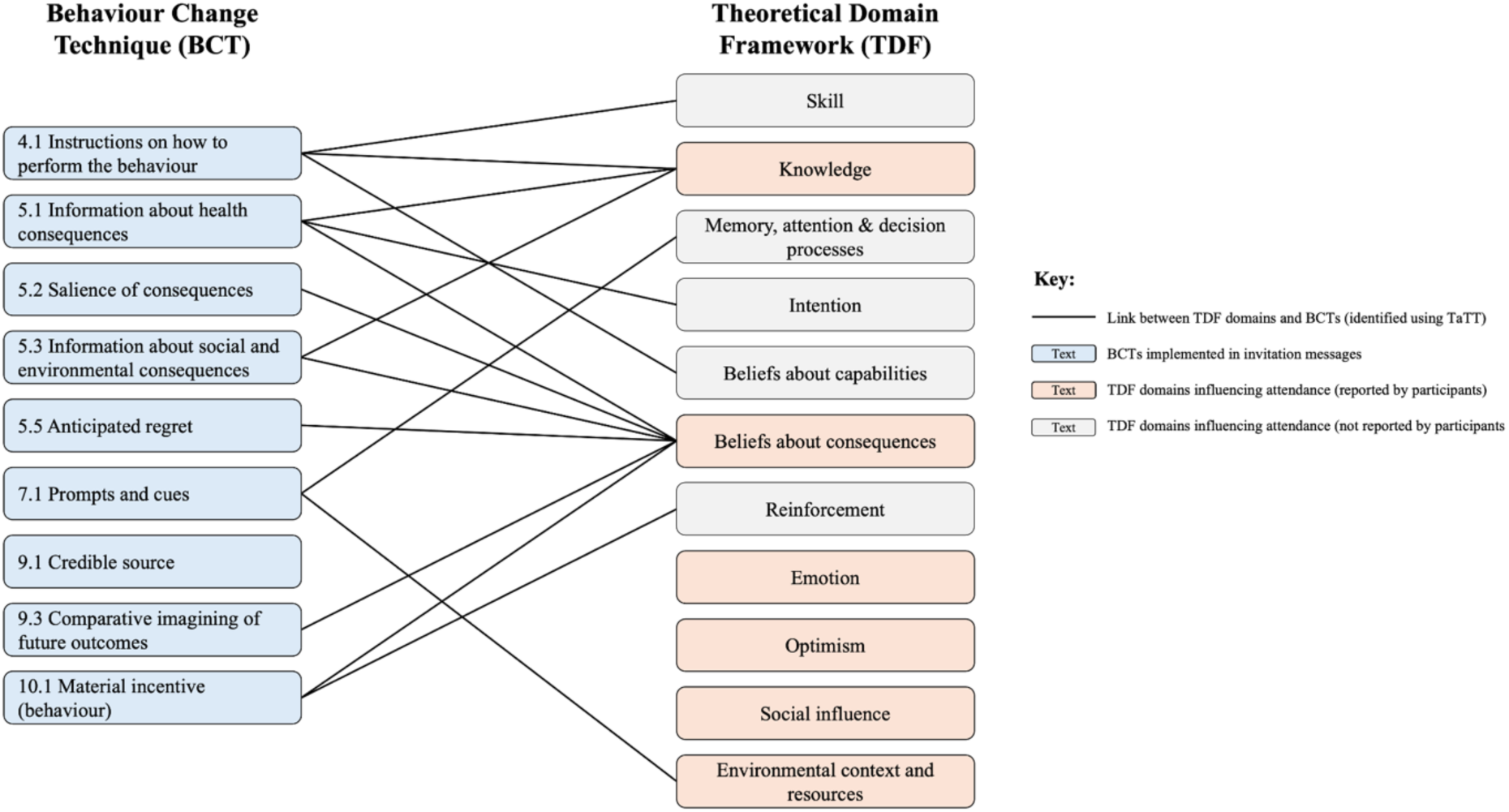
Links between BCTs used in the message conditions and TDF components reported as enablers and barriers by the participants.

All BCTs included in the messages theoretically matched at least one TDF domain, except for Credible Source (9.1). However, three TDF domains that were mapped to participants’ reasons for (non-)attendance, ‘Emotion’, ‘Optimism’, and ‘Social Influence’, were not addressed by any BCTs in the messages. Appendix D contains the results of the BCT coding and which components of the messages represented which BCTs.

In addition, the BCTs coded within the messages were found to address a further five TDF domains that had not been included in the pre-defined list of reasons to (not) attend; Skills, Intention, Beliefs about Capabilities, Reinforcement and Memory, Attention & Decision Processes.

### 3.5 Mediation by feeling scared or nervous

Considering fear as a mediator, the exposure-mediator association was positive: participants in the Messenger condition had 1.65 times higher odds of reporting fear (95% CI [1.02, 2.67], p = .041) against control (Table 5). For other conditions, the estimated odds ratios were smaller, with wide confidence intervals that were not consistent in one direction of association. The adjusted mediator-outcome association was negative: respondents who experienced fear reported, on average, 0.66-points lower likelihood of attending the TLHC (95% CI [-0.87, -0.46], p <.001).

**Table 5.** Exposure-mediator and mediator-outcome associations.

| Exposure → Mediator Models | OR | 95% CI | p |
| --- | --- | --- | --- |
| **Mediator: Feeling Scared** |  |  |  |
| Messenger | 1.65 | [1.02, 2.67] | .041* |

| Gain-frame | 1.42 | [0.87, 2.32] | .163 |
| --- | --- | --- | --- |
| Status quo | 1.29 | [0.78, 2.12] | .321 |
| Challenge | 1.43 | [0.88, 2.34] | .153 |
| **Mediator: Feeling Nervous** |  |  |  |
| Messenger | 1.20 | [0.89, 1.61] | .228 |
| Gain-frame | 0.78 | [0.57, 1.07] | .124 |
| Status quo | 1.21 | [0.90, 1.62] | .204 |
| Challenge | 1.17 | [0.87, 1.57] | .290 |
| **Mediator → Outcome Models** | **β** | **95% CI** | **p** |
| Feeling Scared | -0.66 | [-0.87, -0.46] | <.001* |
| Feeling Nervous | -0.18 | [-0.31, -0.05] | .007* |

*Note. Analyses were based on a sample of 3140 participants. OR = Odds Ratio; β = standardised regression coefficient; CI = confidence interval. Exposure → Mediator models are logistic regressions; Mediator → Outcome models are linear regressions. \*p<.05*

In the models with nervousness as the mediator, the estimated effects of the messaging conditions against control were small, with high imprecision indicated by the wide CIs: the odds ratio for Messenger versus Control was 1.20 (95 % CI [0.89, 1.61], p = .228). The mediator-outcome model found a modest reduction in intention: respondents who felt nervous reported, on average, 0.18-points lower likelihood of attending the TLHC (95 % CI [-0.31, -0.05], p = .007).

The overall effects returned by the mediation analyses were similar in magnitude to the pairwise mean differences in Table 5, which was expected, because the exposure was randomised and no post-assignment confounders were available for inclusion in the adjustment set. Under this specification, the interventional direct (IDE) and interventional indirect effects (IIE) sum to the overall interventional effect (OE), which, in the absence of intermediate confounding, equals the total mean difference.

As noted above, participants in the Messenger condition reported a 0.17-point higher mean likelihood of attending TLHC compared to the Control condition. Most of this difference was captured by the direct effect through pathways other than fear (IDE = 0.19, 95% CI [0.04, 0.34], p = .012) or feeling nervous (IDE = 0.17, 95% CI [0.02, 0.32], p = .025). The results also indicated an opposing indirect path through fear, lowering the overall effect (IIE = -0.01, 95% CI [-0.04, 0.00], p = .064) but this effect was very small and did not reach a conventional significance threshold. The indirect estimate through nervousness was near zero, with a narrow CI (IIE = - 0.00, 95 % CI [-0.01, 0.00], p = .270). For the remaining message conditions, no overall, direct, or indirect effects relative to Control were detected; point estimates were close to zero (Table 6).

**Table 6.** Mediation analysis results with feeling scared and nervous as the mediators.

| Message conditions | Effect | Feeling Scared |  | Feeling Nervous |  |
| --- | --- | --- | --- | --- | --- |
|  |  | Estimate [95% CI] | p | Estimate [95% CI] | p |
| Gain-frame | OE | -0.02 [-0.17, 0.15] | .853 | -0.01 [-0.17, 0.15] | .846 |
|  | IDE | -0.00 [-0.15, 0.16] | .979 | -0.02 [-0.17, 0.15] | .797 |
|  | IIE | -0.02 [-0.03, 0.01] | .209 | 0.00 [-0.00, 0.02] | .165 |
| Messenger | OE | 0.17 [0.02, 0.32] | .028* | 0.17 [0.02, 0.32] | .027* |
|  | IDE | 0.19 [0.04, 0.34] | .012* | 0.17 [0.02, 0.32] | .025* |
|  | IIE | -0.01 [-0.04, 0.00] | .064 | -0.00 [-0.01, 0.00] | .270 |
| Status Quo | OE | -0.02 [-0.19, 0.14] | .803 | -0.02 [-0.19, 0.14] | .798 |
|  | IDE | -0.01 [-0.18, 0.15] | .874 | -0.02 [-0.19, 0.15] | .843 |
|  | IIE | -0.01 [-0.03, 0.01] | .380 | -0.00 [-0.02, 0.00] | .266 |
| Challenge | OE | 0.10 [-0.06, 0.24] | .211 | 0.10 [-0.07, 0.24] | .210 |
|  | IDE | 0.11 [-0.05, 0.25] | .157 | 0.10 [-0.06, 0.24] | .195 |
|  | IIE | -0.00 [-0.03, 0.01] | .200 | -0.00 [-0.01, 0.00] | .322 |

Note. Analyses were based on a sample of 3140 participants. OE = Overall Effect, IDE = Interventional Direct effect, IIE = Interventional Indirect Effect. \* $p < .05$

## 4. Discussion

This study examined 1) whether behaviourally informed invitation messages increased intention to attend the NHS TLHC, 2) identified barriers and enablers to attendance using the TDF, 3) the extent to which invitation messages addressed these influences using the BCW, and 4) the role of fear in shaping intention to attend. Across conditions, behaviourally informed messages performed similarly to the Control, with only the GP-endorsed message (Messenger condition) showing a slightly stronger effect, although this was not robust to adjustment for multiple comparisons. Participants most frequently reported barriers related to *Emotion* (the fear of the screening process and results)*, Social Influence* (perceived judgement and lack of GP recommendation), and *Knowledge* gaps, while the most commonly reported enablers centred around *Beliefs about Consequences* (the perceived benefits of early cancer detection). Mapping of message content to BCTs revealed a degree of misalignment between intervention content and participants’ most frequently reported barriers, as the messages tested primarily targeted *Beliefs about Consequences* and *Knowledge* while largely failing to address the TDF domains of *Emotion* and *Social influences*. Exploratory analyses found that fear and nervousness were associated with lower intention to attend TLHC. The GP-endorsed message seemed to increase reported fear compared to the control message, while fear was negatively associated with intention to attend. These findings align with the broader literature showing inconsistent effects of messaging interventions in cancer screening, particularly low-intensity approaches that do not directly address practical or emotional barriers (Yang et al., 2025; Uy et al., 2017; Sallis et al., 2019).

Misalignment between reported barriers and the BCTs included in the messages could be a reason for the lack of effectiveness of the intervention messages. Only around half of the reported barriers and enablers were addressed by any of the invitation messages (as judged by the theoretical congruence between TDF domains and BCTs), and barriers on *Emotion* and *Social Influence* were not targeted by any BCTs in the messages.

Social influence was the most frequently reported barrier (37.5%), including perceptions of judgement related to smoking status (e.g., “I feel judged for being a smoker or ex-smoker”) and absence of GP endorsement (e.g., “My GP has not recommended it to me”). Feelings of judgement may elicit guilt or shame, consistent with prior research (Pahwa et al., 2025; Olson et al., 2022; Salman et al., 2025), while absence of GP recommendation has been consistently identified as a barrier (Lowenstein et al., 2019; Cavers et al., 2022). Emotional responses also emerged as a key barrier, with 23% reporting feeling scared or nervous, aligning with studies showing fear can deter screening uptake, whether related to a potential cancer diagnosis or the screening process itself (Lowenstein et al., 2019; Schiffelbein et al., 2020; Cavers et al., 2022; Salman et al., 2025). A further 25.8% reported *Knowledge*-related barriers, including misconceptions about treatability (e.g., “I don’t think lung cancer can be treated”) and uncertainty about screening purpose (e.g., “I don’t understand what the screening is for”), also aligned with evidence suggesting insufficient understanding may exacerbate anxiety and reduce perceived relevance (Cavers et al., 2022; Schiffelbein et al., 2020).

In contrast, *Beliefs about Consequences* emerged as a strong enabler, with 81.2% of participants believing that early diagnosis would improve treatment outcomes (e.g. “If cancer is diagnosed earlier, it is more treatable”). This finding aligns with prior research showing that perceived benefits of early detection facilitate screening uptake (Pahwa et al., 2025; Cavers et al., 2022).

Exploratory mediation analyses revealed that the Messenger condition increased fear, with participants who viewed the GP-endorsed message more likely to report feeling scared. Fear, in turn, appeared to act as a barrier: participants who reported feeling scared rated their likelihood of attending lower on average. The modest positive effect of the Messenger condition on intention appeared to operate through other pathways, such as perceived credibility or social influence, with fear neither driving the benefit nor substantially counteracting it. The results suggest that while the Messenger condition may have inadvertently triggered fear, this did not meaningfully diminish its overall effect. Future messages incorporating credible recommendations may nonetheless benefit from explicitly acknowledging and normalising fear, to ensure emotional responses do not undermine otherwise effective content.

This pattern suggests that the Messenger condition’s positive effect on intention operated through unmeasured mechanisms, while simultaneously eliciting fear that, although a barrier to attendance, only negligibly counteracted this benefit. This finding highlights the importance of attending to the emotional consequences of invitation messages. Future iterations might therefore consider explicitly acknowledging and normalising fear while preserving motivating features, such as credible endorsement, so that unmanaged emotional responses do not undermine an otherwise effective message.

One possible explanation for the relative effectiveness of the Messenger condition is that it directly addressed a commonly reported barrier. The Messenger Condition incorporated a *Credible Source* BCT by referencing GP recommendation (e.g. “your GP recommends…”), as shown in Figure 2. This content directly aligns with the Social Influence barrier “My GP has not recommended it to me.” In contrast, other prevalent barriers related to emotion and social influences were not directly targeted by any message content. Although evidence linking *Credible Source* to *Social Influence* is mixed (Carey et al., 2019; Connell et al., 2019), the higher mean intention observed suggests GP endorsement may play a role in addressing social influence barriers in this context.

### 4.1 Implications

The present findings may help explain inconsistent effects observed in the broader literature on messaging-based interventions for cancer screening. There is a lack of alignment between intervention messages and behavioural influences (i.e., the reported barriers). Failure to address these barriers may limit message effectiveness, a pattern that extends to other screening programmes where message design was not informed by prior investigation of population-specific barriers and enablers (e.g. Hagoel et al., 2016; Sallis et al., 2016; Marcotte et al., 2023). The intervention messages in this study were informed by behavioural economics frameworks (Hallsworth et al., 2015), prior NHS Health Check studies (Sallis et al., 2016; 2019), and expert consensus, but lacked a systematic framework for evaluation. Future intervention developments would benefit from following the stages of the BCW, ensuring message content is explicitly mapped to appropriate BCTs informed by empirically identified barriers. For example, emotional barriers such as fear may be addressed through techniques involving social support or reassurance (Lorencatto et al., 2012). A revised invitation message might read: “Attending lung cancer screening may feel worrying, but early checks can help detect problems when treatment is more effective, and your family and loved ones may value you taking this step.”

### 4.2 Strengths and limitations

This study has several strengths. First, we systematically applied behaviour change frameworks to evaluate invitation messages from an NHS screening programme, demonstrating how the application of BCW for intervention evaluation in practice. This contrasts with widely used behavioural economics frameworks, which lack systematic methods for linking behavioural influences on intervention content and evaluation. Second, the study addresses a timely policy priority (House of Commons Library, 2026), providing insights for national rollout. Third, the large sample (n = 3,274) with random allocation strengthens internal validity. Fourth, robust statistical methods were employed to allow valid inference when distributional assumptions are violated.

This study has three main limitations. First, it assessed intention to attend rather than observed attendance. Intentions do not consistently translate into behaviour (Ajzen, 2011; Cane et al., 2012; Conner and Norman, 2022), and the hypothetical message presentation may not fully reflect real-world decision-making (Podsakoff et al., 2003). Future studies should link invitation messaging to attendance data. Second, the control message contained more information than current TLHC invitation letters, limiting direct comparison of intervention effects and the applicability of findings to practice. Third, the mediation analyses were exploratory and should be interpreted cautiously. Randomisation strengthens causal interpretation of message effects on the mediators but does not protect the mediator-outcome relationship from confounding. Although we adjusted for available sociodemographic and health-related covariates, unmeasured influences such as pre-existing health anxiety may have affected both the mediators and intention. The simultaneous measurement of mediators and outcome further limits causal interpretation.

## 5. Conclusion

The behaviourally informed messages tested were not substantially more effective than the NHS invitation-based control message in increasing intention to attend screening, though tentative evidence suggested the GP-endorsed message may confer a small advantage, this effect was not robust to multiple comparison adjustment and was modest at best.

Analysis using the TDF demonstrated that participants’ decisions were shaped primarily by emotional responses, particularly fear, and social influences such as perceived judgement and absence of GP endorsement, with beliefs about early detection benefits acting as a key enabler. Application of the BCW revealed that invitation messages only partially addressed these influences, with emotional and social barriers largely unaddressed by the BCTs used. Exploratory analyses indicated fear was modestly associated with lower intention to attend; the GP-endorsed message appeared to increase fear compared to the standard message, but without clear evidence this undermined the message’s overall impact. Future message design should seek to decouple credible recommendation from fear elicitation by addressing emotional concerns directly. These findings highlight the importance of systematically aligning message content with empirically identified behavioural influences when designing interventions to improve lung cancer screening uptake.

## Data Availability

All data produced are available online at OSF: https://osf.io/qvjwy/

https://github.com/xiaozhou-tan/TLHC_Scripts

## List of Abbreviations

ANOVA: Analysis of Variance
BCT: Behaviour Change Technique
BCTTv1: Behaviour Change Technique Taxonomy version 1
BCW: Behaviour Change Wheel
CI: Confidence Interval
IDE: Interventional Direct Effect
IIE: Interventional Indirect Effect
LCS: Lung Cancer Screening
LDCT: Low-dose Computed Tomography
MINDSPACE: Messenger, Incentives, Norms, Defaults, Salience, Priming, Affect, Commitment, Ego
NHS: National Health Service
OE: Overall Effect
SD: Standard Deviation
TaTT: Theory and Techniques Tool
TDF: Theoretical Domains Framework
TLHC: Targeted Lung Health Check

## Declarations

### Ethical approval and consent to participate

Please see the methods section for information regarding ethical approval and data protection.

### Consent for publication

All authors have seen and approved the final version of this manuscript for publication.

### Availability of data and materials

The data used in this study is publicly available on the Open Science Framework (OSF) repository at https://osf.io/qvjwy/files/osfstorage/663cf46d419d00400cfea245.

## Author’s contribution

Conceptualisation: XT, LP, CG

Data Curation: XT, MD, TME, SU

Investigation: TME, SU, PK

Methodology: XT, MD, LP, CG

Formal Analysis: XT, MD, LP, CG

Resources: TME

Software: XT, MD

Supervision: MD, LP, CG, TME, SJ

Validation: MD, LP, CG

Writing – Original Draft Preparation: XT

Writing – Review & Editing: XT, MD, LP, CG, TME, SJ, SU, PK

## Acknowledgements

The authors would like to thank Charlotte Smith, Project Manager for Early Diagnosis (Targeted Lung Health Checks), NHS Cancer Programme, NHS England, and Poppy Richards, Programme Manager for Early Diagnosis Programmes (Lung), NHS Cancer Programme, NHS England

## Supplementary Materials

The R scripts used to generate the results and figures are available in the GitHub repository at https://github.com/xiaozhou-tan/TLHC_Scripts.

## Appendix A

### Overview of the original NHS England online experiment

#### Aims

The original NHS England Behavioural Science Unit experiment was designed to generate rapid evidence on whether small changes to the naming and wording used in communications could improve understanding of, and intentions to engage with, the Targeted Lung Health Check (TLHC) programme. Specifically, the experiment aimed to: (1) test whether alternative programme names and initial appointment names improve comprehension of what the service is and what the first appointment involves; (2) assess whether alternative names are perceived as clearer and more likely to encourage participation; (3) test whether different invitation-style messages influence stated likelihood of taking part; and (4) explore participants’ emotional responses to the messages and self-reported reasons for wanting to attend or not attend.

#### Design

The study was an online randomised experiment embedded within a survey. Eligibility criteria for participant recruitment included being aged 55 to 74 years, residing in England, and having a history of smoking (current or former smokers).

Eligible participants were randomly allocated to see:

- One of five programme names (e.g., the current name “Targeted lung health check” versus alternatives containing the term “lung cancer screening”).
- One of five initial appointment names (e.g., the current “lung health check” versus alternatives such as “initial lung cancer risk assessment”).
- One of five invitation messages, each presented after a common explanatory paragraph describing what the initial screening appointment involves (duration/format and that a healthcare professional asks questions to assess lung cancer risk).

The primary outcomes were:

1. Comprehension: whether participants correctly identified (from multiple-choice options) the purpose of the programme and what the initial appointment involves.
2. Perceived clarity and perceived likelihood to take part: ratings of each name (after participants were later shown all candidate names).
3. Likelihood to take part after the message: rated after exposure to one of the message variants.
4. Affective responses to the message (participants selected up to three emotions from a list) and motivations/barriers (participants selected up to three reasons for wanting to attend and up to three reasons for not wanting to attend).

### Measures and Procedures

Participants were recruited via a market research agency (Bilendi) and NHS England panels (NHS App panel and NHS Vaccines panel). Eligibility criteria were applied via screening questions: participants were aged 55–74, based in England, and current or former smokers, matching the core eligibility group for the programme. Recruitment aimed to oversample smokers and groups that had been more difficult to engage in prior work (including ethnic minority groups and lower socioeconomic groups). Data collection took place in October 2023.

#### Programme name comprehension and perceptions

Participants were randomly assigned to view one of five names for the overall screening programme. Programme name comprehension was assessed using a multiple-choice item asking participants what they believed the programme was for. Perceived impact of the name on attendance was assessed using self-report ratings of how likely each name would be to make them attend screening.

#### Appointment name comprehension and perceptions

Participants were shown one of five names for the initial screening appointment. Comprehension was assessed via a multiple-choice item asking what participants believed the appointment would involve. Participants also rated how likely the appointment name would be to make them attend screening.

#### Invitation message responses

Participants were randomly allocated to one of five invitation messages encouraging attendance at a Targeted Lung Health Check appointment. Following message exposure, participants rated their likelihood of attending on a seven-point scale and selected up to three emotions describing how the message made them feel from a predefined list.

#### Reasons for attending or not attending

Participants selected up to three reasons for wanting to attend and up to three reasons for not wanting to attend a Targeted Lung Health Check from predefined lists. These items captured perceived enablers and barriers to screening participation.

#### Demographic and health-related measures

Participants provided demographic and health-related information, including age, gender, socioeconomic indicators, smoking status, and the presence of long-term health conditions.

## Appendix B

### Message 1: Control

The NHS recommends that people aged 55 to 74 years old who have ever smoked has lung cancer screening. This is a short appointment that can take place in person, by phone or online. A healthcare professional will ask you some questions about your medical history and lifestyle to work out your risk of having lung cancer. *We are working with NHS England to offer FREE Lung Cancer Screening and would like to invite you to arrange an appointment*.

### Message 2: Gain-frame (experiential/emotive)

The NHS recommends that people aged 55 to 74 years old who have ever smoked have lung cancer screening. This is a short appointment that can take place in person, by phone or online. A healthcare professional will ask you some questions about your medical history and your lifestyle to work out your risk of having lung cancer.

Lung cancer causes more deaths than any other cancer in the UK. Lung cancer screening works out how healthy your lungs are. It aims to find any signs of lung cancer at an early stage, when it is usually easier to treat, and treatment is more successful. Make sure you can keep doing the things you love and living your life. Book a lung cancer screening appointment to stay healthy.

### Message 3: Messenger effect

The NHS recommends that people aged 55 to 74 years old who have ever smoked have lung cancer screening. This is a short appointment that can take place in person, by phone or online. A healthcare professional will ask you some questions about your medical history and your lifestyle to work out your risk of having lung cancer.

Based on your age and smoking history, your GP recommends you have lung cancer screening. If you have lung cancer, you are three times more likely to find it early if you go for lung cancer screening. Early treatment can often cure lung cancer, giving you the best possible chance to go on to live a healthy life.

### Message 4: Status quo

The NHS recommends that people aged 55 to 74 years old who have ever smoked have lung cancer screening. *Lung cancer screening is are now available in your area and it is your turn to book your appointment.* This is a short appointment that can take place in person, by phone or online. A healthcare professional will ask you some questions about your medical history and your lifestyle to work out your risk of having lung cancer. *Don’t delay, confirm your appointment right away*.

### Message 5: Challenging assumptions (I don’t want to bother the NHS)

The NHS recommends that people aged 55 to 74 years old who have ever smoked have lung cancer screening. This is a short appointment that can take place in person, by phone or online. A healthcare professional will ask you some questions about your medical history and your lifestyle to work out your risk of having lung cancer.

Screening can find signs of lung cancer early. This can help you get the treatment you need sooner, often with more successful results. If you wait until your health worsens, you can cost the NHS more money to treat you, and you are less likely to get better.

## Appendix C

### Full list of reasons to and not to attend the NHS Targeted Lung Health Check and their Theoretical Domains Framework (TDF) coding

Participants were asked to select up to three items each for reasons to attend and reasons not to attend the NHS Targeted Lung Health Check. CG, LP, and XT independently reviewed the response options and coded them using the Theoretical Domains Framework (TDF; Cane et al., 2012). Tables B1 and B2 present the full list of items for reasons to attend and not to attend, respectively, along with their corresponding TDF classifications.

Participants’ reported enablers and barriers (i.e., reasons to attend and reasons not to attend, respectively) were grouped by TDF components, as multiple items mapped to the same domain. Frequencies were then calculated separately for enablers and barriers, with the aggregated results presented in Table B3.

**Table C1.** TDF coding for reasons to attend.

*TDF coding for reasons to attend*
| Reasons to attend | TDF components |
| --- | --- |
| If cancer is diagnosed earlier it is more treatable | Beliefs about consequences |
| If my GP recommended me to | Social influence |
| My family would want me to | Social influence |
| I always attend health screening when invited | Not coded* |
| If I have cancer it would be better for me to find out at an earlier stage | Beliefs about consequences |
| It is something I can do to take control of my health | Optimism |
| It would be reassuring to get a negative result | Emotion |
| It is free | Environmental context and resources |

\*Not enough information to make a coding decision without making assumptions.

**Table C2.** TDF coding for reasons not to attend.

*TDF coding for reasons not to attend*
| Reasons not to attend | TDF components |
| --- | --- |
| My GP has not recommended it to me | Social influence |
| I would be scared of a cancer diagnosis | Emotion |
| I would not want to know if I have lung cancer | Not coded* |
| I don't think lung cancer can be treated | Beliefs about consequences |
| I don't have any symptoms | Knowledge |
| I feel judged for being a smoker/ex-smoker | Social influence<sup>§</sup> |
| I don't think I am risk of lung cancer | Believes about consequences |
| I am nervous about the screening process | Emotion |
| I don't think lung screening is worthwhile | Beliefs about consequences |
| I don't understand what the screening is for | Knowledge |
| It is expensive or difficult to attend | Environmental context and resources |

\*Not enough information to make a coding decision without making assumptions.
<sup>§</sup>Though “smoker/ex-smoker” could also fit into ‘Identity’, but ‘Identity’ cannot be a barrier to attending TLHC.

**Table C3.**
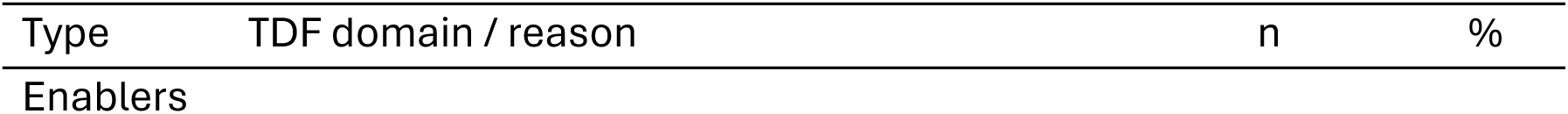
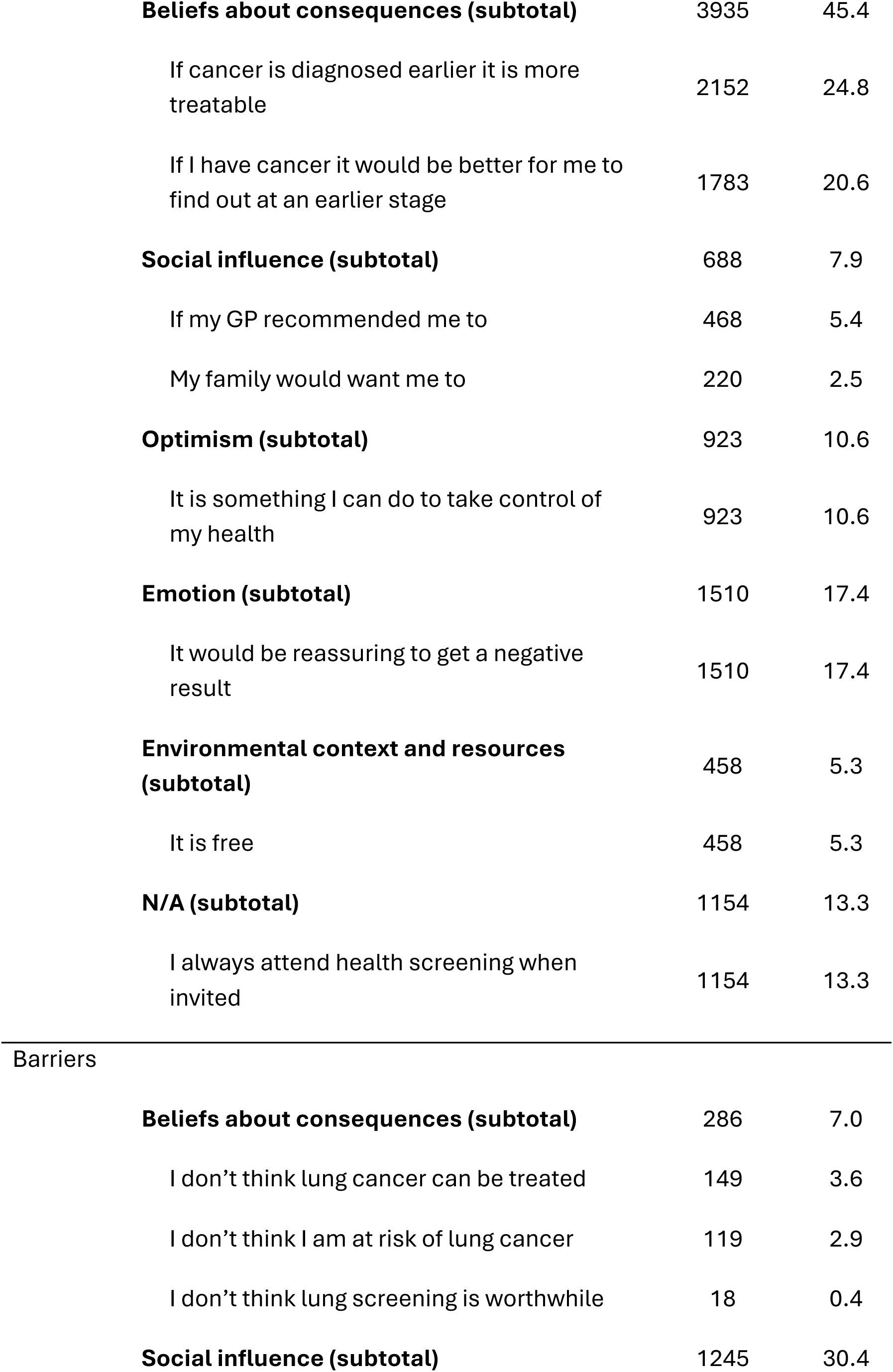
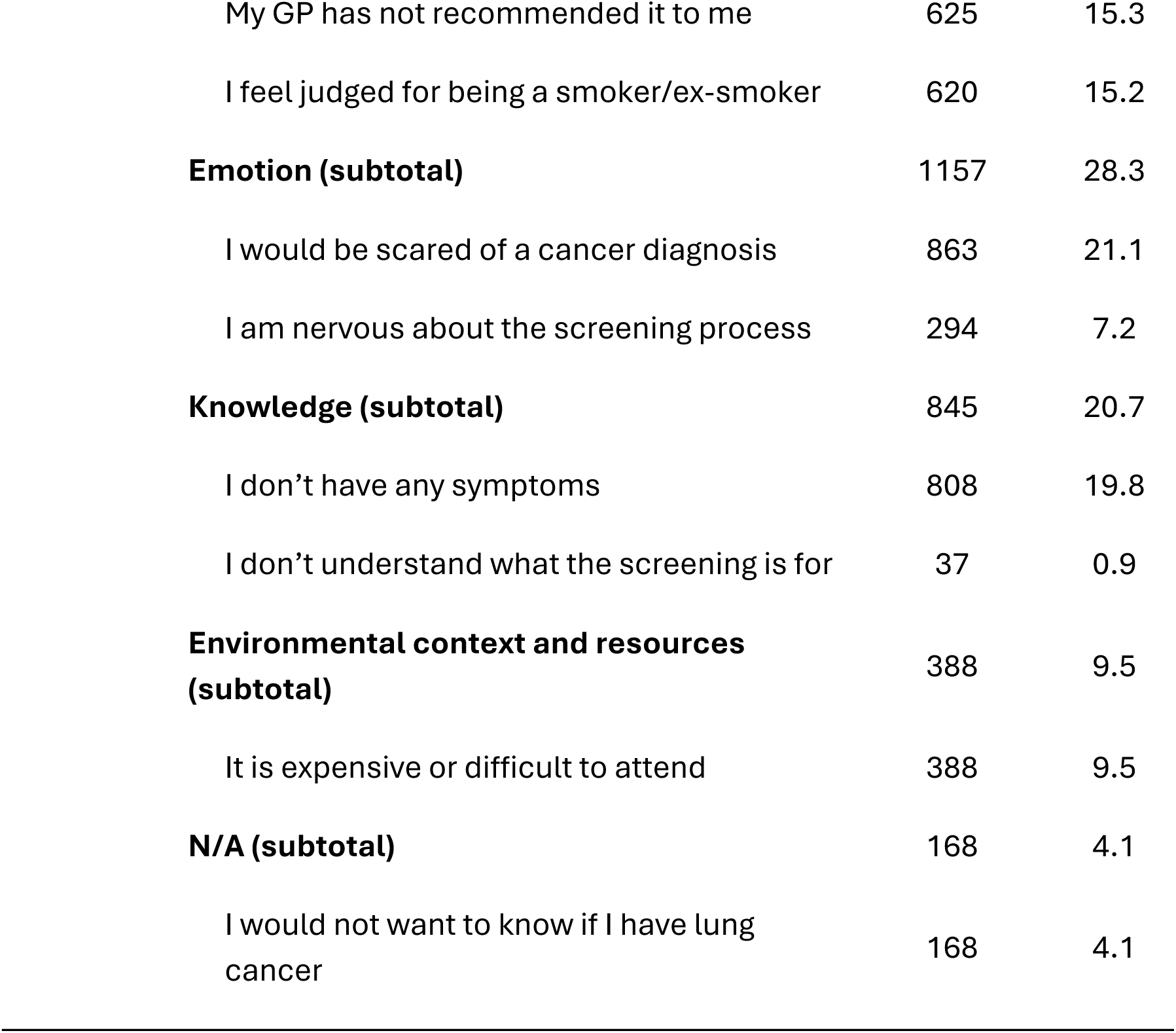
Frequencies of TDF components reported by the participants.

| Type | TDF domain / reason | n | % |
| --- | --- | --- | --- |
| Enablers |  |  |  |

|  |  |  |
| --- | --- | --- |
| **Beliefs about consequences (subtotal)** | 3935 | 45.4 |
| If cancer is diagnosed earlier it is more treatable | 2152 | 24.8 |
| If I have cancer it would be better for me to find out at an earlier stage | 1783 | 20.6 |
| **Social influence (subtotal)** | 688 | 7.9 |
| If my GP recommended me to | 468 | 5.4 |
| My family would want me to | 220 | 2.5 |
| **Optimism (subtotal)** | 923 | 10.6 |
| It is something I can do to take control of my health | 923 | 10.6 |
| **Emotion (subtotal)** | 1510 | 17.4 |
| It would be reassuring to get a negative result | 1510 | 17.4 |
| **Environmental context and resources (subtotal)** | 458 | 5.3 |
| It is free | 458 | 5.3 |
| **N/A (subtotal)** | 1154 | 13.3 |
| I always attend health screening when invited | 1154 | 13.3 |

Barriers
|  |  |  |
| --- | --- | --- |
| **Beliefs about consequences (subtotal)** | 286 | 7.0 |
| I don't think lung cancer can be treated | 149 | 3.6 |
| I don't think I am at risk of lung cancer | 119 | 2.9 |
| I don't think lung screening is worthwhile | 18 | 0.4 |
| **Social influence (subtotal)** | 1245 | 30.4 |
| My GP has not recommended it to me | 625 | 15.3 |
| I feel judged for being a smoker/ex-smoker | 620 | 15.2 |
| **Emotion (subtotal)** | **1157** | **28.3** |
| I would be scared of a cancer diagnosis | 863 | 21.1 |
| I am nervous about the screening process | 294 | 7.2 |
| **Knowledge (subtotal)** | **845** | **20.7** |
| I don't have any symptoms | 808 | 19.8 |
| I don't understand what the screening is for | 37 | 0.9 |
| **Environmental context and resources (subtotal)** | **388** | **9.5** |
| It is expensive or difficult to attend | 388 | 9.5 |
| **N/A (subtotal)** | **168** | **4.1** |
| I would not want to know if I have lung cancer | 168 | 4.1 |

## Appendix D

### Behaviour Change Techniques (BCTs) identified in invitation messages for each message condition

Participants were randomly assigned to one of five message conditions. CG, LP, and XT reviewed the message content and identified Behaviour Change Techniques (BCTs) present within each condition, coding them qualitatively using the Behaviour Change Techniques Taxonomy v1 (BCTTv1; Michie et al., 2013). Table D summarises the BCTs assigned to each message condition and the corresponding message content. Messages in all conditions shared a same common paragraph, which is coded with the Control condition in Table D:

> “The NHS recommends that people aged 55 to 74 years old who have ever smoked have lung cancer screening. This is a short appointment that can take place in person, by phone or online. A healthcare professional will ask you some questions about your medical history and lifestyle to work out your risk of having lung cancer.”

**Table D.**
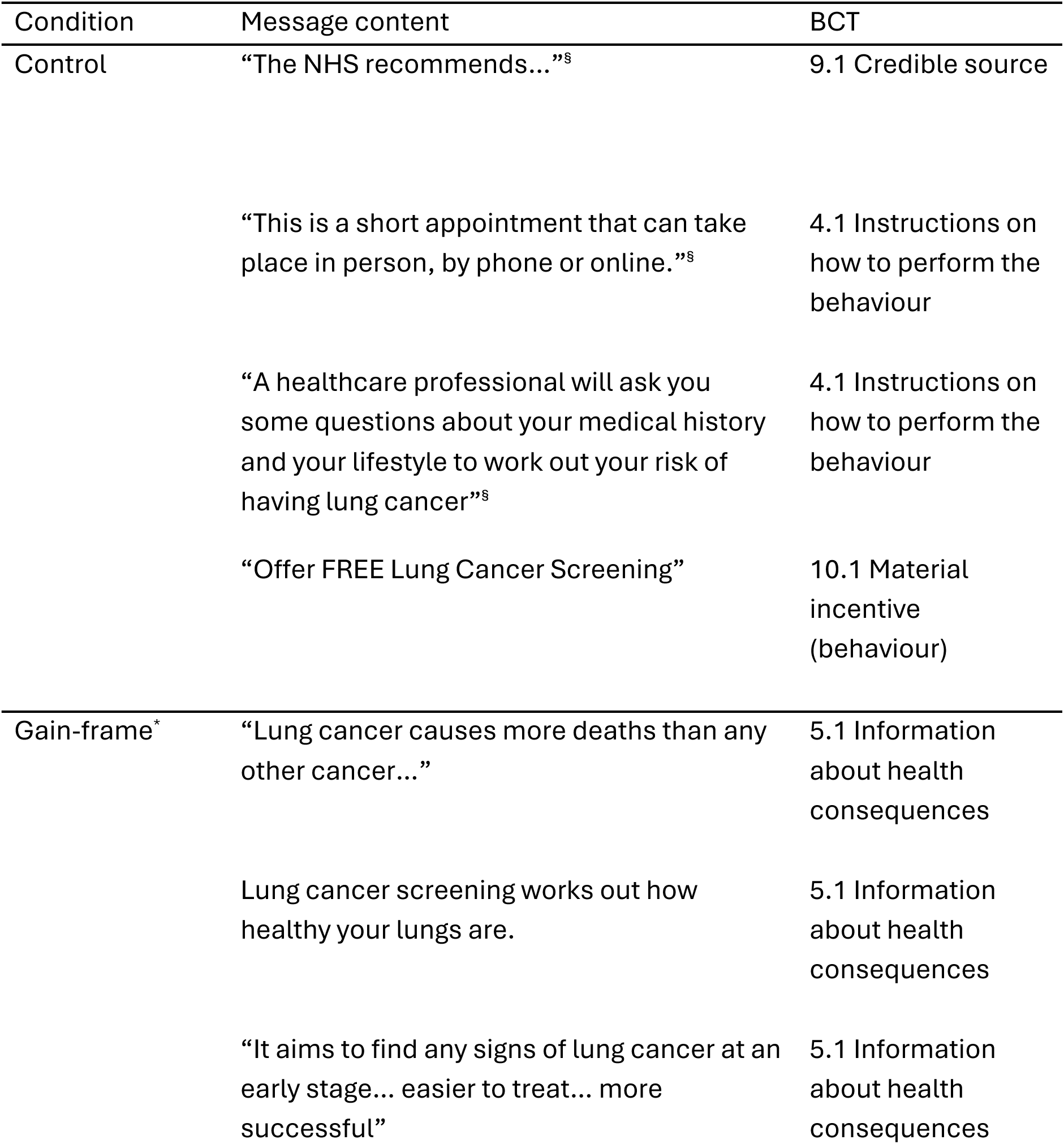
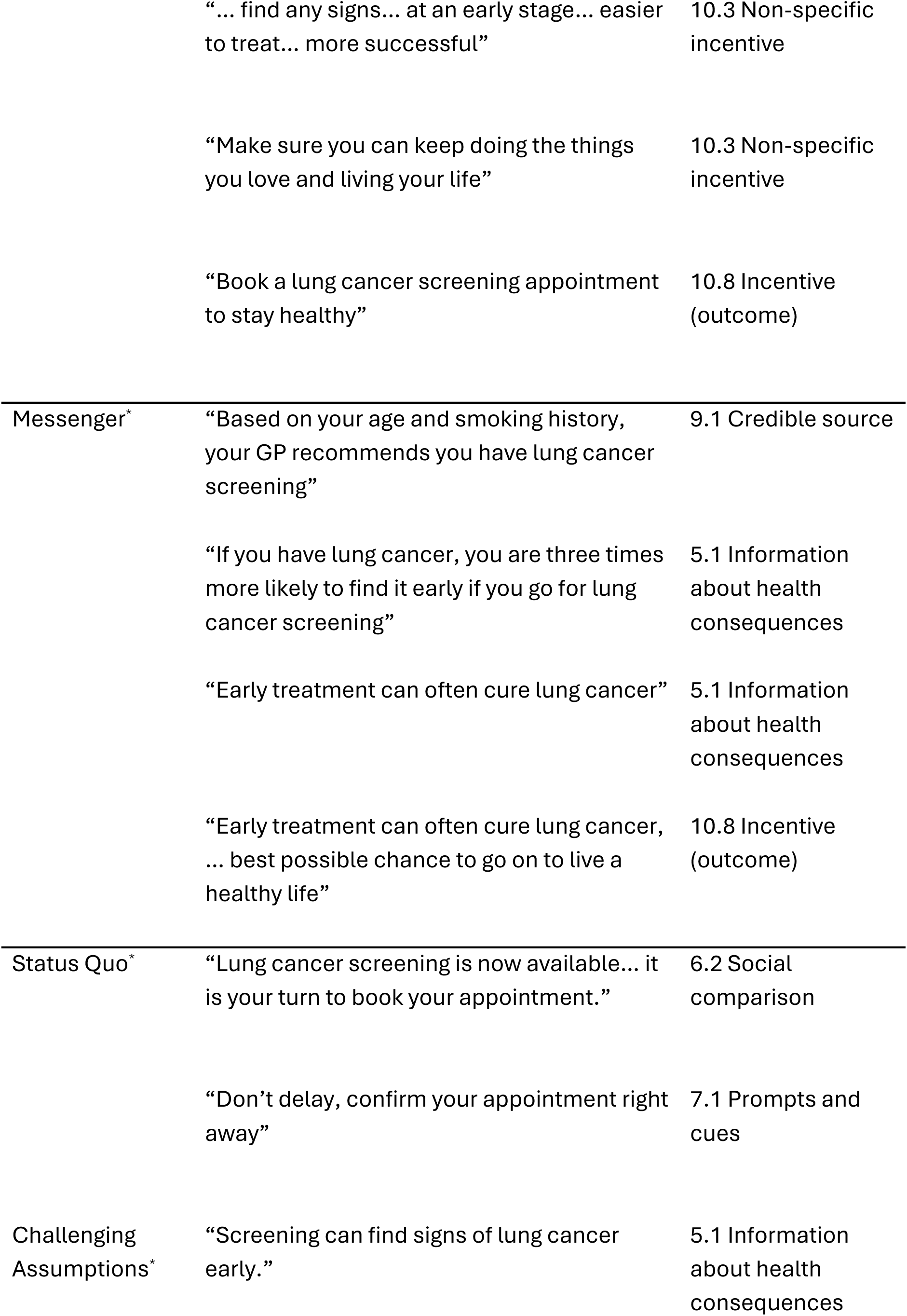
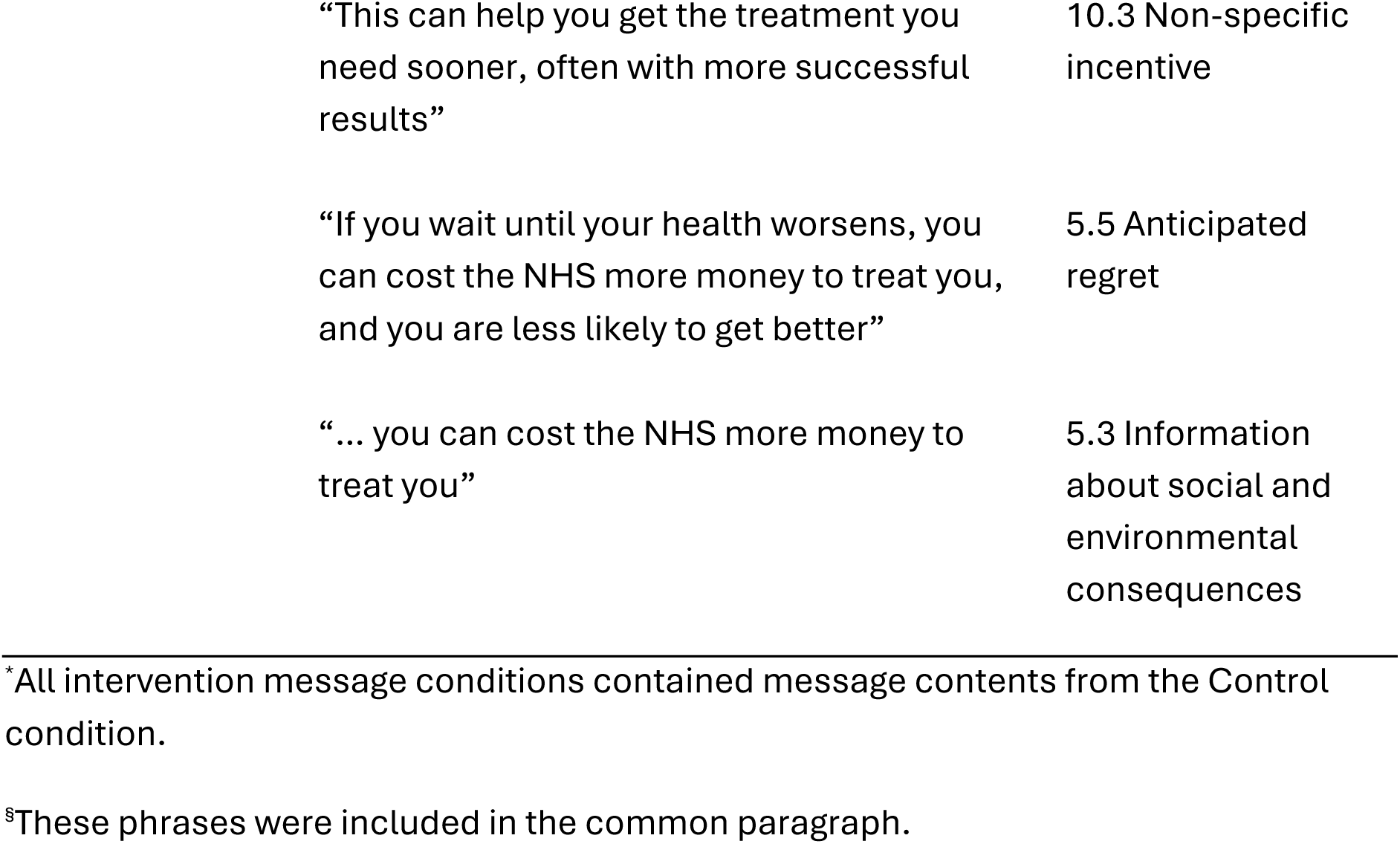
Behaviour Change Techniques identified across message conditions.

| Condition | Message content | BCT |
| --- | --- | --- |
| Control | “The NHS recommends...”<sup>s</sup> | 9.1 Credible source |
|  | “This is a short appointment that can take place in person, by phone or online.”<sup>s</sup> | 4.1 Instructions on how to perform the behaviour |
|  | “A healthcare professional will ask you some questions about your medical history and your lifestyle to work out your risk of having lung cancer”<sup>s</sup> | 4.1 Instructions on how to perform the behaviour |
|  | “Offer FREE Lung Cancer Screening” | 10.1 Material incentive (behaviour) |
| Gain-frame* | “Lung cancer causes more deaths than any other cancer...” | 5.1 Information about health consequences |
|  | Lung cancer screening works out how healthy your lungs are. | 5.1 Information about health consequences |
|  | “It aims to find any signs of lung cancer at an early stage... easier to treat... more successful” | 5.1 Information about health consequences |
|  | "... find any signs... at an early stage... easier to treat... more successful" | 10.3 Non-specific incentive |
|  | "Make sure you can keep doing the things you love and living your life" | 10.3 Non-specific incentive |
|  | "Book a lung cancer screening appointment to stay healthy" | 10.8 Incentive (outcome) |
| Messenger* | "Based on your age and smoking history, your GP recommends you have lung cancer screening" | 9.1 Credible source |
|  | "If you have lung cancer, you are three times more likely to find it early if you go for lung cancer screening" | 5.1 Information about health consequences |
|  | "Early treatment can often cure lung cancer" | 5.1 Information about health consequences |
|  | "Early treatment can often cure lung cancer, ... best possible chance to go on to live a healthy life" | 10.8 Incentive (outcome) |
| Status Quo* | "Lung cancer screening is now available... it is your turn to book your appointment." | 6.2 Social comparison |
|  | "Don't delay, confirm your appointment right away" | 7.1 Prompts and cues |
| Challenging Assumptions* | "Screening can find signs of lung cancer early." | 5.1 Information about health consequences |

|  |  |
| --- | --- |
| “This can help you get the treatment you need sooner, often with more successful results” | 10.3 Non-specific incentive |
| “If you wait until your health worsens, you can cost the NHS more money to treat you, and you are less likely to get better” | 5.5 Anticipated regret |
| “... you can cost the NHS more money to treat you” | 5.3 Information about social and environmental consequences |

\*All intervention message conditions contained message contents from the Control condition.
§These phrases were included in the common paragraph.

